# Prior exposure to antiretroviral therapy among adult patients presenting for HIV treatment initiation or re-initiation in sub- Saharan Africa: a systematic review

**DOI:** 10.1101/2022.10.19.22281280

**Authors:** Mariet Benade, Mhairi Maskew, Allison Juntunen, Sydney Rosen

## Abstract

**Background:** As countries in sub-Saharan Africa have scaled up access to antiretroviral therapy (ART) for HIV, patient attrition rates of up to 30% per year have created a large pool of individuals who initiate treatment with prior ART experience (non-naïve re-initiators). Little is known about the proportion of non-naïve re-initiators within the population presenting for treatment initiation.

**Methods:** We conducted a comprehensive, rapid review of recent peer-reviewed reports that presented data on proportions of adult patients initiating ART who were treatment naïve and non-naive in sub-Saharan Africa. Searching PubMed, EMBASE, Web of Science, and international conference abstracts, we sought studies published after 1 January 2018 with data collected after January 2016, when universal HIV treatment access became the norm. We included clinical trials and observational studies and accepted self-report, laboratory discernment of antiretroviral metabolites, or viral suppression at initiation or previously reported in the medical record as evidence of prior exposure. We report results of each eligible study and identify gaps in the literature.

**Results:** Of 1,782 articles returned in our initial search, we found nine, describing ten cohorts, that contained sufficient information for the review, of which half were from South Africa. The proportion of the study samples with evidence of prior ART use ranged from 5% (self-report only) to 53% (presence of ART metabolites in hair or blood sample among self-reported naïve patients). The vast majority of studies that were screened did not report proportions of initiators who were non-naïve, and among the few that did, the metrics used to determine and report non-naïve proportions were inconsistent and difficult to interpret.

**Conclusion:** The proportion of patients initiating HIV treatment who are truly ART-naïve is not well documented in the literature. From the studies identified, it seems likely that 20% to 50%—and likely at least 30%—of ART patients who present for ART are re-initiators. Standard reporting metrics and diligence in reporting this characteristic of ART initiation cohorts are needed, as is research to understand the reluctance of patients to report prior ART exposure.

## Introduction

The successful scale-up of access to antiretroviral therapy (ART) for HIV treatment in sub-Saharan Africa has produced a growing population of patients who have interrupted or stopped treatment sometime since they started, either permanently or temporarily. While very recent numbers on attrition from ART programs are scarce, retention in care rates for the region were reported to average 78% at 12 months after treatment initiation in a review published in 2015, suggesting that for a cohort of patients initiating in any given year, nearly a quarter have been lost from care one year later[1]. Many of these lost patients, however, proceed to “re-initiate” treatment in the months or years after dropping out of treatment programs[2]. Two estimates posit the extent of treatment re-initiation. The first, from the U.S. President’s Emergency Plan for AIDS Relief (PEPFAR) reported that more than 580,000 patients returned to care after a treatment interruption in just the quarter from July to September 2020 in the countries that PEPFAR supports[3]. The second was from the Western Cape Province of South Africa, where among the subset of patients whose CD4 counts were less than 50 cells/mm^3^, the proportion with prior treatment experience rose from 14% to 57% between 2008 and 2017[4].

Outside of indirect estimates such as those mentioned above, little is known about the actual proportions of non-naïve patients among all those presenting for ART initiation. Accurate data are difficult to obtain, largely because most HIV medical record systems neither distinguish between naïve and re-initiators nor allow tracking from one healthcare facility to another or over long intervals of inactivity. In most countries, a patient who originally initiated ART at one facility and then dropped out of care can easily present as a new patient at a nearby facility and be assumed to be ART-naïve. Self-reported information about a patient’s naïve or non-naïve status may be unreliable, because patients who are known to have stopped treatment may be reprimanded, provided poorer service by healthcare facility staff, or required to participate in multiple adherence training sessions, creating an incentive to present oneself as a new patient regardless of prior experience[5].

As the number of re-initiators continues to increase, understanding the proportion and characteristics of ART initiators who are not treatment-naïve is an important step in improving overall HIV treatment outcomes. By definition, treatment re-initiators previously faced barriers to retention in care that they were unable to overcome. Common barriers to retention include logistical challenges such as transport costs, psychosocial deterrents such as stigma, and personal preferences[6,7], and these barriers may become more prohibitive for patients who have already withdrawn from care once. Achieving long-term retention in care targets may thus require that healthcare systems differentiate interventions and services for re-initiators from those offered to naïve initiators. Re-initiators also comprise an increasing proportion of patients presenting with advanced HIV disease[4], who require additional care beyond simple ART initiation.

As national treatment programs mature, the proportion of ART initiators who are non-naïve will continue to grow, making the few available earlier estimates obsolete. To help fill the gap in empirical evidence on current proportions of naïve and non-naïve ART initiators, we conducted a comprehensive, rapid review of recently published or presented (2018 and later) peer-reviewed reports in sub-Saharan Africa that directly or indirectly presented data on re-initiation rates.

## Methods

Following World Health Organization guidance for rapid reviews[8] we conducted a rapid systematic review of peer-reviewed publications and conference abstracts that reported on prior exposure to antiretroviral therapy among adult patients presenting for HIV treatment in sub-Saharan Africa. The review was registered on the International Prospective Register of Systematic Reviews (PROSPERO CRD42022324136).

### Search strategy, study selection, and data extraction

For this review, our primary outcome was the proportion of adults presenting for ART initiation (initially or after interruption) in public sector HIV treatment programs in sub-Saharan Africa who are not ART naïve. To indicate prior ART use, we accepted self-reported questionnaire responses, rates of viral load suppression at ART initiation (suggesting previous ART use), medical record evidence (e.g. a prior viral load test), and biological measurements of the presence of antiretroviral metabolites in blood, hair, or urine specimens at or prior to treatment initiation. Inclusion and exclusion criteria for the review are shown in S1 Table.

To identify potential sources, we developed a search string with the assistance of a medical librarian. The search syntax included variations of the terms HIV, treatment, antiretroviral therapy, retention, and adherence, limited to sub-Saharan Africa. During the course of the review, we made three revisions to the original protocol as submitted to PROSPERO, in order to maximize the potential for finding relevant sources. Specifically, we 1) included studies focusing on pregnant women starting ART in PMTCT programs; 2) added “undisclosed” and “retention” as search terms, as shown in S2 Table; and 3) included studies that required a minimum duration of ART for enrollment, as explained below. Search strings can be found in S2 Table.

We searched the PubMed, Embase, and Web of Science databases and abstracts for the International AIDS Conference, International AIDS Society (IAS) Conference on HIV Science, and Conference on Retroviruses and Opportunistic Infections (CROI) with a search string developed to identify English-language publications which reported on HIV treatment initiation in sub-Saharan Africa from 1 January 2018 until 31 March 2022. We further limited our search to articles from which the majority of the data were generated in 2016 or later, as this was when universal treatment access became common in the region. Follow-up searches extended the latest publication or conference presentation date of the review to September 15, 2022.

We included cohort, cross-sectional, case-control, and interventional studies that reported primary data on initiation of ART in the adult (≥ 18 years old) population, including for PMTCT. We included indexed pre-prints but excluded unpublished reports. We also excluded commentaries, modeling studies, and other sources that did not report primary data. While we excluded systematic reviews and meta-analyses, we manually searched existing systematic reviews for additional, non-duplicate references to be included. Where more than one publication reported on the same patient cohort, we chose the one that was either most recent or provided the most relevant data.

Because we were interested specifically in reports of the proportion of patients presenting for ART initiation or re-initiation in routine care who were naïve and non-naïve, we excluded studies that stated that prior ART experience was an exclusion criterion for the study, with one exception. If a study included only participants who self-reported as naïve but were then found to have evidence of previous ARV exposure, we included that proportion as a result, accepting that it explicitly omits patients who excluded themselves because of prior exposure and thus almost certainly reflects an underestimate of the true population prevalence of previous exposure. We also included studies that only enrolled patients who had achieved a specified duration of follow up on ART—whether one month or several years—despite the fact that these studies would have missed patients who were lost from care prior to reaching that specified duration.

All peer reviewed references identified using the respective search strings from PubMed, Embase and Web of Science were imported into Rayyan QCRI, where deduplication occurred. An initial, independent, blinded review (reviewers were not aware of each other’s decisions) of the titles and abstracts was conducted by three study team members (MB, AJ, SR) using Rayyan QCRI. A full text review was then conducted for all publications remaining after the initial review by two study team members (MB, AJ or MB, SR), with conflicts resolved through discussion. Reasons for excluding publications were recorded during the full text review. As a quality check, one author (SR) also checked a sample (10%) of the excluded sources against exclusion criteria. At each stage of the review process any conflicts between reviewers were assessed and resolved though consensus of three authors (MB, AJ, SR). The results of the search were documented in accordance with the PRISMA-P reporting checklist (S3 Table).

We created a data extraction tool to capture study and sample characteristics, proportions with previous ART exposure, and the type of indicator of previous exposure reported (e.g. self-report, laboratory results).

### Outcomes and analysis

Our outcome of interest was the proportion of ART initiates who were treatment naïve at ART initiation, defined as a patient presenting for initiation of ART who has never previously taken antiretroviral therapy for treatment of HIV (“new initiator”), compared the proportion who were treatment-experienced at ART initiation, defined as a patient presenting for initiation of ART who had previously taken antiretroviral therapy for HIV treatment but had interrupted that therapy for a minimum of 3 months (“re-initiator”). We accepted each paper’s source of information about participants’ status: self-report, medical record review, viral suppression, or laboratory tests for ARV metabolites and report that source in our results.

To evaluate the data, we first report each paper’s outcome, with descriptive information regarding the population and setting to which the results apply. As described in the Prospero protocol, we had intended to estimate pooled results for individual countries and populations and to stratify by patient and facility characteristics, but we ultimately identified too few eligible sources to allow for any pooled or stratified analysis.

Quality of the eligible studies was assessed using the Joanna Briggs Institute Critical Appraisal Checklists[9].

## Results

### Sources identified

The results of the systematic search are shown in Figure 1. A total of 1,782 non-duplicate abstracts of peer reviewed journal articles and 9 abstracts from the selected conferences were screened. After the initial title and abstract review, 1,450 articles and abstracts were excluded, leaving 332 documents for full-text review. During the full review, an additional 322 were excluded. Reasons for exclusions are reported in S4 Table. The primary reason for exclusion was lack of information on the naïve or non-naïve status of participants.

**Figure 1.**
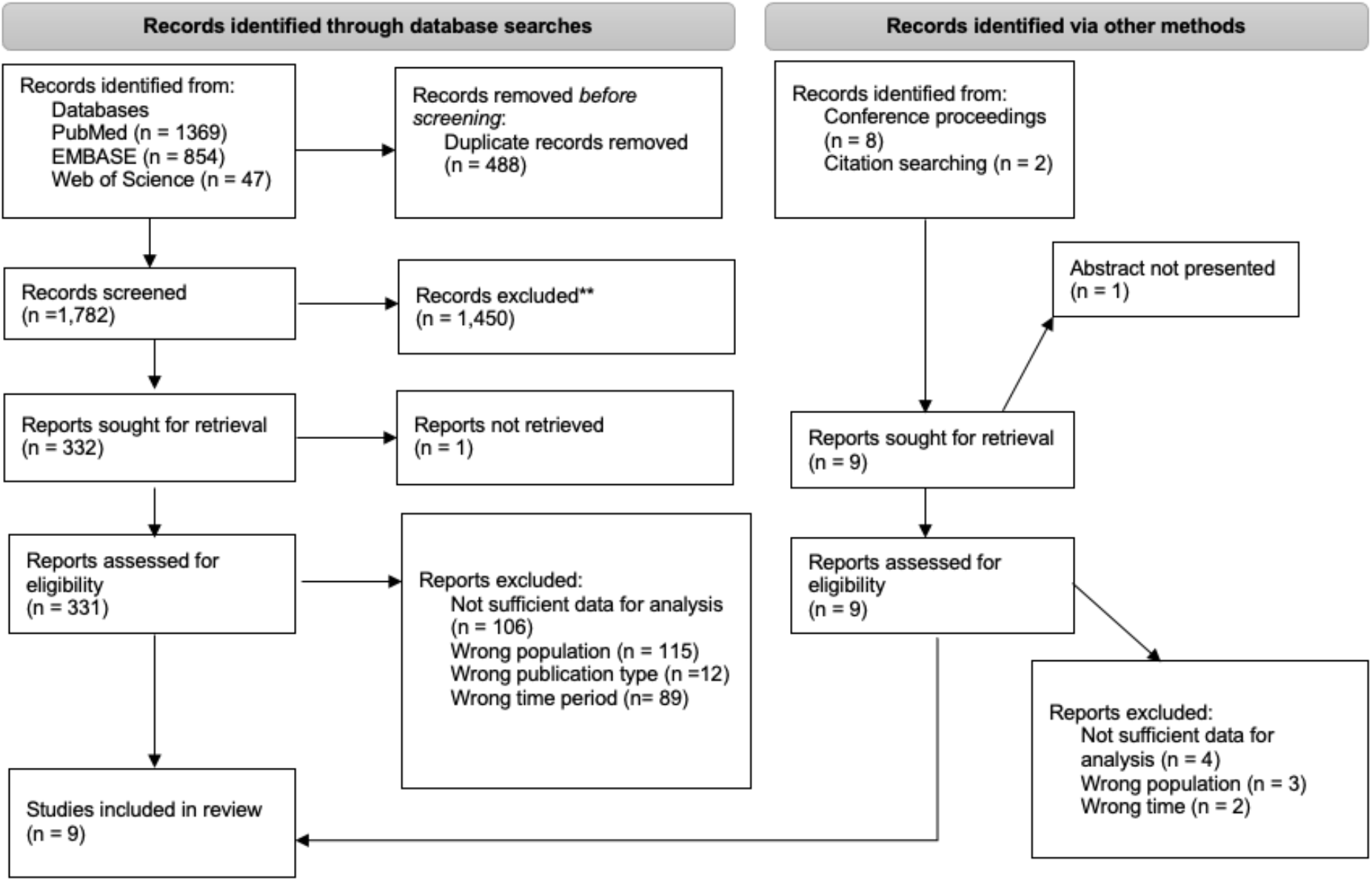
PRISMA flow chart.

Nine peer-reviewed articles were retained in the final data set for the full review, including one that reported data from two countries and will be included in our analysis as two studies, creating a total of ten sets of results. The studies are described in Table 1.

**Table 1.** Studies included in the review.

| Source<br>(alphabetical<br>by country) | Country<br>and<br>location | Study design<br>and source of<br>data | Study<br>population | Dates of<br>presentation<br>for ART<br>initiation | Sample<br>size | Sex (%<br>female) | Age<br>(median,<br>IQR) |
| --- | --- | --- | --- | --- | --- | --- | --- |
| Lebelonyane<br>2020[10] | Botswana<br>(national<br>sample) | Subset of<br>baseline data<br>for<br>intervention<br>arm of cluster<br>randomized<br>trial of HIV<br>prevention | Adults found to<br>be HIV-positive<br>during<br>community HIV<br>prevention<br>campaign and<br>linked to HIV<br>care | June 2016-March 2018 | 800 | 55% | 33 (26-41) |
| Buju<br>2022[11] | Democratic<br>Republic of<br>Congo<br>(Bunia) | Observational<br>prospective<br>cohort of<br>patients<br>receiving<br>dolutegravir | Adults<br>presenting for<br>ART initiation or<br>ongoing ART<br>treatment at all<br>ART clinics in<br>Busia | July 2019-July<br>2021 for those<br>initiating<br>during the<br>study; earlier<br>for those<br>already on ART | 177 | 69% | 39 (12)* |
| Genet 2021<br>[12] | Ethiopia<br>(Northwest<br>Ethiopia) | Observational,<br>cross sectional<br>study of first<br>line treatment<br>failure | Adults† who<br>had received at<br>least 6 months<br>of ART | May October<br>2017 | 430 | 58% | 38 (12-67)** |
| Kunzweiler<br>2018 [13] | Kenya<br>(Kisumu) | Observational<br>prospective<br>cohort | Adult men who<br>have sex with<br>men (MSM) | August 2015-September<br>2016 | 63 | 0% | 27 (22-32) |
| Rosen<br>2019[14] | Kenya<br>(Kericho,<br>Kapsabet,<br>and<br>Kombewa<br>counties) | Baseline data<br>for<br>intervention<br>arm of clinical<br>trial of same-<br>day ART<br>initiation | Adults<br>presenting for<br>ART initiation at<br>3 public sector<br>hospitals | July 2017-April<br>2018 | 477 | 58% | 36 (29-44) |
| Dorward<br>2020[15] | South<br>Africa<br>(KwaZulu<br>Natal<br>Province) | Baseline data<br>for randomized<br>controlled trial<br>for point-of-<br>care HIV viral<br>load testing | Adults clinically<br>stable on ART<br>and due for 6-month viral load<br>testing | August 2016-February 2017<br>(est) | 390 | 60% | 32 (27-38) |
| Maskew<br>2020[16] | South<br>Africa<br>(Gauteng<br>Province) | Baseline data<br>for<br>intervention<br>arm in clinical<br>trial of same-<br>day ART<br>initiation | Adults<br>presenting for<br>ART initiation at<br>3 public sector<br>clinics | March-September<br>2018 | 296 | 64% | 35 (30-44) |
| Mavhandu-Ramarumo<br>2019[17] | South<br>Africa<br>(Limpopo<br>Province) | Baseline<br>samples from<br>clinical trial of<br>drug resistance | Adults<br>presenting for<br>ART initiation at<br>3 public sector<br>clinics | 2017-2019 | 77 | 90% | 35 (27-42) |
| Rosen 2019<br>[14] | South<br>Africa<br>(Gauteng<br>Province) | Baseline data<br>for<br>intervention<br>arm of clinical<br>trial of same-<br>day ART<br>initiation | Adults<br>presenting for<br>ART initiation at<br>3 public sector<br>clinics | March-July<br>2017 | 600 | 63% | 34 (29-41) |
| Sithole<br>2021[18] | South<br>Africa<br>(KwaZulu<br>Natal<br>Province) | Subset of<br>baseline data<br>from clinical<br>trial of home-<br>based ART<br>initiation | Non-pregnant<br>adults<br>presenting for<br>ART initiation at<br>2 public sector<br>clinics, with CD4<br>>100 and no<br>active TB | February 2018-November<br>2018 | 193 | 60% | Not<br>reported |

\*Mean (standard deviation)
†Study included children under 18, but they comprised <7% of the study sample
\*\*Mean (minimum-maximum)

The first five studies in Table 1 were conducted in Botswana, the Democratic Republic of Congo (DRC), Ethiopia and Kenya, and the remaining five all in South Africa. Seven of the ten reported baseline data from a clinical trial conducted for other purposes, while three were observational studies. All were very or somewhat small in size, enrolled adult patients presenting or having previously presented for routine ART initiation at public sector clinics, and collected most or all data prior to the disruption in service delivery caused by COVID-19 in early 2020[19]. Specific populations enrolled varied by study, though only one, Kunzweiler 2018, was limited to a non-general adult population.

### Proportions of patients non-naïve

In Table 2, we report the proportion of patients in each study reported to be non-naïve when presenting for ART initiation.

**Table 2.** Proportions of cohorts reported to be non-naïve at ART initiation.

| Source | Source of data | Proportion non-naïve at initiation | Inclusion criteria related to prior ART exposure | Comments |
| --- | --- | --- | --- | --- |
| Lebelonyane 2020* | Not stated; only indicated as participants who had “previous ART treatment” | 54/800 (7%) | Sample included 16-17 year olds; minors may be less likely than adults to have had an opportunity for prior ART exposure. | Participants were identified through community-based testing and referred for ART; sample does not represent routine walk-in ART initiation population. |
| Buju 2022 | Viral load suppressed at ART initiation | 93/177 (52%) | None. | Enrolled both initiators patients and patients already on ART; results presented are only for initiators. |
| Genet 2021 | Self-report | 90/430 (21%) | Sample included 12-17 year olds; minors may be less likely than adults to have had an opportunity for prior ART exposure. Participants required to have completed 6 months on ART; patients lost to follow up before reaching 6 months were excluded. | Enrolled patients younger than 18. Those on second line ART were excluded; patients eligible for second line treatment may be more likely to be re-initiators. |
| Kunzweiler 2018 | Viral load suppressed at ART initiation | 19/63 (30%) (13 of 19 with suppressed viral load at ART initiation self-reported being ART-naïve) | Excluded 9 participants who were lost to follow up before reaching 6 months on ART. | Also excluded 3 patients who did not have viral load test results. |
| Rosen 2019 | Self-report | 18/240 (8%) | Included self-reported re-initiators who had interrupted ART for ≥ 3 months. | Patients who self-reported that they had previously initiated ART but had stopped for ≥3 months were eligible; those who self-reported that they had interrupted for < 3 months were excluded. |
| Dorward 2020 | Self-report | 18/390 (5%) | Participants required to have completed 6 months on ART; patients lost to follow up before reaching 6 months were excluded. |  |
| Maskew 2020 | Self-report | 33/296 (11%) | Included self-reported re-initiators who had interrupted ART for ≥ 3 months | Patients who self-reported that they had previously initiated ART but had stopped for ≥3 months were eligible; those who self-reported that they had interrupted for < 3 months were excluded. |
| Mavhandu-Ramarumo 2019 | Laboratory assay of blood and/or hair sample for presence of TDF, EFV, and/or FTC metabolites | 41/77 (53%) | All participants self-reported as ART naïve | Stated enrollment criteria included naïvete; those who had been on ART previously may have self-screened out of this study. |
| Rosen 2019 | Self-report | 7/298 (2%) | Included self-reported re-initiators who had interrupted ART for ≥ 3 months | Patients who self-reported that they had previously initiated ART but had stopped for ≥3 months were eligible; those who self-reported that they had interrupted for < 3 months were excluded. |
| Sithole 2021<sup>†</sup> | Undetectable viral load at ART initiation | 62/193 (32%) (Other outcomes reported: 42/193 (22%) had medical record evidence of prior ART use; 37/193 (19%) had detectable ARV metabolites in blood.) | All participants self-reported as ART naïve | Stated enrollment criteria included naïvete; those who had been on ART previously may have self-screened out of this study. Participants were identified through community-based testing; sample does not represent routine walk-in ART initiation population. Primary DO-ART trial excluded 588 participants out of 2,479 who were virally suppressed at time of initiation and 66 who were found to already be on ART[20] |

TDF, Tenofovir; EFV, Efavirenz; FTC, Emtricitabine
\*A separate publication by the same author team, reporting data that overlapped with those reported in this study, stated that “22% of advanced HIV patients had previously been on ART.” This sample was limited to those with AHD, however.
<sup>†</sup>The parent study on which this study was based.

The proportion of patients presenting for ART initiation who were reported to be non-naïve ranged from 2 to 53%. It is important to note that the results shown in Table 2 are not strictly comparable to one another, however, due to the different data sources, populations, and relevant exclusions listed in the right-hand column of the table. In Maskew 2021 and Rosen 2020, non-naïve patients were allowed to enroll as long as they had been off ART for at least three months. Nevertheless, these studies, which are the only ones that relied entirely on participant self-report, reported the lowest proportion of non-naïve participants. Two other studies in Table 2, Mavhandu-Ramarumo 2019 and Sithole 2021, explicitly excluded self-reported non-naïve patients; all patients enrolled in these studies claimed not to have been on ART previously. The relatively high proportions of non-naïve patients in these studies are thus still minimum estimates of the true proportion of ART initiators with prior treatment exposure at the study clinics, as anyone who admitted to prior use of ARVs will have been excluded from study enrolment. Similarly, Kunzweiler 2018 and Dorward 2020 excluded patients lost to follow up before 6 months after ART initiation, and thus may have underestimated the proportion re-initiating if non-naïve patients are more likely than naïve patients to drop out of care. Study participants were also drawn from diverse populations of individuals with HIV, including those identified in a community campaign (Lebelonyane 2020), in a household survey as part of a randomized trial (Sithole 2021), or as routine, walk-in presenters at clinics (Buju 2022, Maskew 2020, Rosen 2019).

Because all the eligible studies were small and/or had primary outcomes other than the proportion naive, little stratification of results by facility or patient characteristics was reported. Mavhandu-Ramarumo 2019 reported that 7 of the 8 (88%) males in the sample population had evidence of prior ART exposure, compared to only 49% of the 34 females. In contrast, Sithole 2021 found that women (37%) were more likely to have evidence of undisclosed ART use than men (25%). Other characteristics that were associated with undisclosed ART use by undetectable viral load were younger age (35% in 18-29 year olds, 30% in 30-49 year olds, and 23% among those >50 years) and living with a partner who was HIV positive (44% compared to 37%; adj OR 1.94 (95% CI 0.95-3.96)). Kunzweiler 2018’s results apply specifically to MSM. Other studies did not provide information for stratification by facility or patient characteristics.

### Quality of evidence

Each of the studies included in the review either presented baseline enrollment data from a randomized controlled trial or observational data. Outcomes after ART initiation were not relevant for our review, which looked only at the status of patients at baseline (ART initiation). For Maskew 2020 and Rosen 2019, data on self-reported naïve status were limited to the intervention arm. The setting, inclusion criteria, and baseline characteristics for these studies were clearly defined. Mavhandu-Ramarumo 2019, Lebelonyane 2020, Dorward 2020, and Sithole 2021 also sufficiently defined inclusion criteria, study setting, and baseline characteristics.

We assessed Kunzweiler 2018, Genet 2021, and Buju 2022 as observational studies. Kunzweiler 2018 utilised snowball sampling, as is frequently done among key populations at high risk of stigma. All three papers described the study setting and sample population of interest clearly, including age, sex and clinical characteristics of HIV presentation.

As is indicated in Table 2 and discussed further below, each of the studies included in this review used a different indicator of prior ART exposure (non-naivete), and most had limitations as to their accuracy and/or the representativeness of their populations.

## Discussion

We systematically reviewed peer-reviewed evidence on the proportions of patients presenting for treatment initiation in sub-Saharan Africa who are or are not ART-naïve. The proportions non-naïve in the sources we found ranged from a low of 2% using self-report only to a high of 53% based on a laboratory analysis of ARV metabolites in blood and hair samples of patients who self-reported to be naïve.

Perhaps the most striking finding of this review is the sheer lack of published evidence to answer our research question. Despite a comprehensive search of the literature published between 2018 and 2022, and including data since 2016, we identified only nine sources and ten cohorts that reported this information, and most included it in only in passing. Half the studies were conducted in South Africa and were relatively small in size; those from other countries provided very little detail. Based on the published and presented research alone, it is fair to say that very little is known about the true proportion of ART initiators who are not treatment-naïve in South Africa, and almost nothing is known about the rest of the region or about specific sub-populations or risk groups. While it is possible that more information is available to program managers who have access to routinely collected medical record data, nothing in the literature suggests that such information is being generated on a large scale or, more important, utilized for program improvement.

During our search, we made a concerted effort to find additional eligible sources, in the hope that there would be more data to review and analyze. This included adding additional search terms, including data on pregnant and post-partum women, and reviewing reference lists from relevant systematic reviews. We reviewed an unusually large number of full-text manuscripts in the hope that they would include proportions naïve and non-naïve in their cohort descriptions (typically “Table 1”) even though there was no indication of this in the abstract.

Unfortunately, most of the sources that originally appeared promising were found to be ineligible, for various reasons. Most simply did not report on baseline naïve/non-naïve status at ART initiation, using any indicator. Some intentionally excluded non-naïve patients prior to enrolment and then reported all participants as naïve without further investigation. For studies that explicitly screened out non-naïve participants, we considered calculating proportions non-naive based on reported numbers of potential participants included and excluded, but we realized that many studies only applied the non-naïve criterion after screening potential participants out for other reasons. We thus could not safely rely on numbers screened out due to non-naïve status for our numerator and had to exclude those papers. Even some of the papers we did include either provided only a passing reference about prior ART exposure, making us uncertain that we interpreted them correctly (e.g. Lebelonyane 2020), or explicitly included patients lost to follow up in the early treatment period (e.g. Kunzweiler 2018 and Dorward 2020).

The small number of eligible papers we did find offer some useful information. They used several different indicators for identifying non-naïve patients, and the indicators produced results that are consistent with their expected accuracy. Maskew 2020 and Rosen 2019, which relied solely on self-report, and Lebelonyane 2020, for with the source of data is unknown, found the lowest proportions non-naïve. Sithole 2021, which excluded a priori anyone self-reporting prior utilization, reported that 32% of patients had evidence of prior use based on being virally suppressed. Buju 2022 and Kunzweiler 2018 reported that 52% and 30%, respectively, of patients presenting for initiation already had suppressed viral loads, suggesting prior ARV use. Finally, Mavhandu-Ramarumo 2019, which also excluded a priori anyone admitting prior utilization, using the most rigorous methodology with both blood and hair samples, estimated 53% of patients had prior ART exposure. The large observed difference between males and females and the small number of males in this study, however, suggest caution in applying the results to male patients.

Based on results of the studies that used laboratory tests, we assume that those that relied on self-report—e.g. Maskew 2020, Rosen 2019, and Dorward 2020—underestimated the true proportion of participants who were non-naïve at initiation. While study inclusion criteria may have biased the samples in Sithole 2021 and Mavhandu-Ramarumo 2019, these, together with Buju 2022, Kunzweiler 2018, and Genet 2021, suggest that it is reasonable (and conservative) to conclude that between 20% and 50% of ART patients—and likely at least 30%--who present for ART are re-initiators. This proportion can be expected to increase with each passing year, as the number of truly naïve HIV-positive individuals declines. If this is so, then re-initiators comprise an important sub-population whose needs are likely to differ from those of naïve initiators and to whom service delivery should be tailored.

As is evident from the discussion above, this review had several limitations. First, while we believe that our search of the peer-reviewed, published literature and abstracts was thorough, the lack of standard terminology for describing prior ART exposure hampered the creation of precise search strings, and it is possible that some sources were missed. Second, we found information from only four of sub-Saharan Africa’s 46 countries, and five of our ten observations were from a single country, South Africa. Since each country in sub-Saharan Africa has a different experience with attrition from ART and approach to re-initiation, results may not be generalizable. Third, as explained in the introduction, even such data as are available tend to be incomplete, due to the limitations of self-reporting and of existing medical record systems.

Fourth, the wide range of results identified may reflect study methodologies, but it may also indicate substantial geographic diversity in outcomes that we cannot address with the data available. We speculate that additional relevant data are collected by program managers, ministries of health, and others but are either not analyzed to answer our research question or simply not published and therefore not accessible. Fifth, the small number of eligible sources, small sample sizes, and heterogeneity of research methods made it impossible to aggregate the results or produce meaningful summary statistics, beyond the range discussed above. Sixth, recall bias may be present in the studies that asked participants after six months or more on ART to self-report their naïve/non-naïve status at the time they initiated treatment. Finally, the fact that six of ten cohorts included were from clinical trials that provided patient compensation may have biased enrollments, though we cannot know how this may have affected patient enrolment or self-report of prior ART usage and the subsequent direction of the bias.

In addition to the sheer dearth of information available to answer our question, the search reported here revealed three important research priorities. First, there is a need for a standard terminology to describe patients with prior ART exposure and prior ART initiation experience. Even the binary terms “naïve” and “non-naïve” can be unclear if patients have previously used antiretroviral medications for PrEP or prevention of vertical transmission. Terms such as “ART experienced” and “ART exposed” are often substituted for non-naïve, without specification of what they refer to. Prior ART use may be “disclosed” or “self-reported.” Similarly, the duration of treatment interruption that leads to “re-initiation” is rarely specified, and “re-initiation” and “re-engagement” are used interchangeably. We can assume that patient returning to care after an interruption of less than one month is not likely to be regarded as a re-initiator, and a patient returning after an interruption of more than one year will likely be defined as a re-initiator. But what of patients with interruptions of 6 or 8 months? A common terminology for describing the phenomenon addressed here would be of great assistance in understanding its magnitude.

Second, in view of the potentially very high proportion of re-initiators among “new” ART patients, it is critical that researchers begin to report proportions naïve and non-naïve as a standard variable when describing patient cohorts, even if data come solely from self-report. We cannot determine from the literature whether many studies do collect this information but omit it from their reports or if it has simply not been collected. We identified several papers that came close to indicating a proportion non-naïve but did not explain their findings clearly enough to include in this review, suggesting that many studies do indeed have access to the relevant information. The study mentioned in our introduction from South Africa’s Western Cape Province, for example, provided detailed information about patients with advanced HIV disease and suggested that more than a third of all patients with very low CD4 cell counts had previously been on ART but were now off, but we could not calculate the overall proportion of non-naïve initiators from the data reported [4]. In earlier years, when the proportion of non-naïve patients was low because treatment programs were still rapidly expanding, the question of prior ART experience may not have been a priority. In view of the results of the few studies available, it is clearly a priority now.

Finally, the phenomenon of large numbers of patients who decline to reveal prior ART use, even when asked directly, is concerning. For studies that intend to limit participation to naïve patients, stated exclusion of those who are non-naïve may encourage non-disclosure, to avoid being denied study enrolment on this basis. Even so, it appears likely that many patients opt to lie about prior exposure. Other research suggests that they have good reason for doing so, as clinics may refuse to re-initiate those who admit to prior default and/or may provide poorer service to them[21–23]. Creating a clinic and community atmosphere that promotes honesty about prior exposure should also be a priority.

In conclusion, while we recognize that simply knowing the proportion of non-naïve patients in a given population will not in itself improve the quality of service delivery, measuring the size of the problem is a critical step in creating momentum to development and implement interventions targeted specifically at re-initiators. Since these are patients who have already demonstrated that they face obstacles to remaining in care, identifying and targeting them for appropriate services is a vital step in improving the outcomes of treatment programs.

## Data Availability

All data used in this study have previously been published and citations are provided.

## Authors’ contributions

MM, SR, and MB conceived of and designed the study. MB, SR, and AJ identified and reviewed sources and extracted data. MB and SR analyzed the data and drafted the manuscript. All authors reviewed and edited the manuscript.

## Additional files

**S1 Table.** Inclusion/exclusion criteria for publications and abstracts

| Parameter | Inclusion criteria | Exclusion criteria |
| --- | --- | --- |
| Population | Ages 18+ years; confirmed HIV positive status; presenting for initiation of any regimen of lifelong antiretroviral treatment | Paediatric and adolescent populations aged <18 years; currently only receiving ART for HIV prevention (PEP or PrEP) |
| Geographic region | Sub-Saharan Africa | None |
| Intervention | None, observational descriptive outcome | None |
| Study design | Reports primary, patient-level data from retrospective or prospective cohorts collected under any study design (trial, observational) with or without a comparison group; systematic reviews, meta-analyses | Case series or reports, purely qualitative studies, treatment guidelines, mathematical models, editorials, commentaries, study or trial protocols |
| Required descriptive data | Describes all of patients, location, timing of ART initiation, facility type, service delivery models and services provided to the public sector through government-managed public health infrastructure or through NGO/private programs or facilities that serve the uninsured sector | Insufficient description of the characteristics needed to describe the study population and outcome |
| Comparator | Not required; single arm evaluations are eligible | None |
| Outcomes | Reports proportion of patients initiating ART that are ART naïve and proportions of patients previously experienced on ART for any duration after initiation. | Insufficient detail provided to estimate of outcome |
| Timing | A majority of data collected for ART initiation on or after January 1, 2016 | A majority of data accrued before January 1, 2016 |

**S2 Table.**
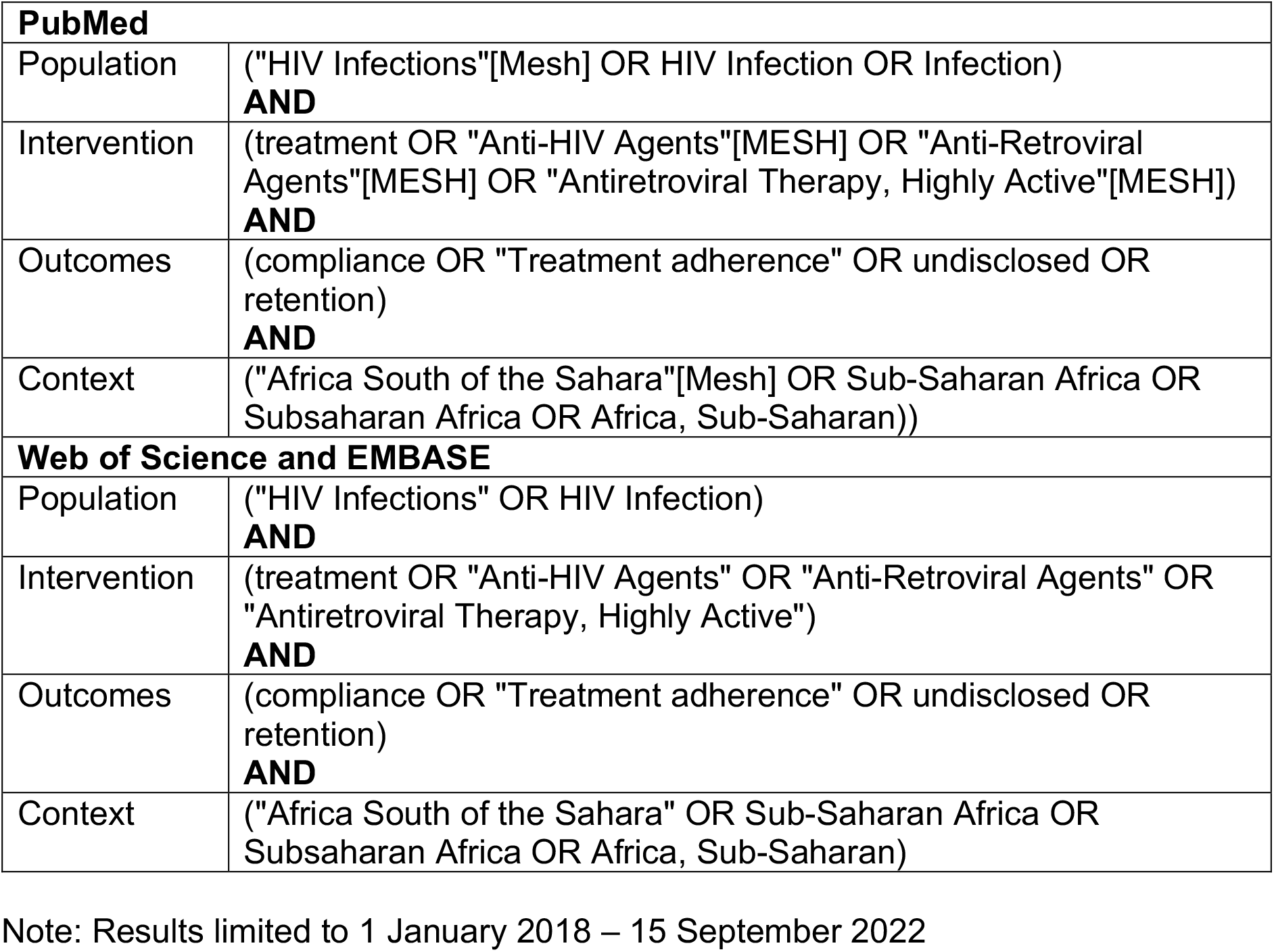
Search strategy

| **PubMed** |  |
| --- | --- |
| Population | ("HIV Infections"[Mesh] OR HIV Infection OR Infection)<br>**AND** |
| Intervention | (treatment OR "Anti-HIV Agents"[MESH] OR "Anti-Retroviral Agents"[MESH] OR "Antiretroviral Therapy, Highly Active"[MESH])<br>**AND** |
| Outcomes | (compliance OR "Treatment adherence" OR undisclosed OR retention)<br>**AND** |
| Context | ("Africa South of the Sahara"[Mesh] OR Sub-Saharan Africa OR Subsaharan Africa OR Africa, Sub-Saharan)) |
| **Web of Science and EMBASE** |  |
| Population | ("HIV Infections" OR HIV Infection)<br>**AND** |
| Intervention | (treatment OR "Anti-HIV Agents" OR "Anti-Retroviral Agents" OR "Antiretroviral Therapy, Highly Active")<br>**AND** |
| Outcomes | (compliance OR "Treatment adherence" OR undisclosed OR retention)<br>**AND** |
| Context | ("Africa South of the Sahara" OR Sub-Saharan Africa OR Subsaharan Africa OR Africa, Sub-Saharan)) |

Note: Results limited to 1 January 2018 – 15 September 2022

**S3 Table.**
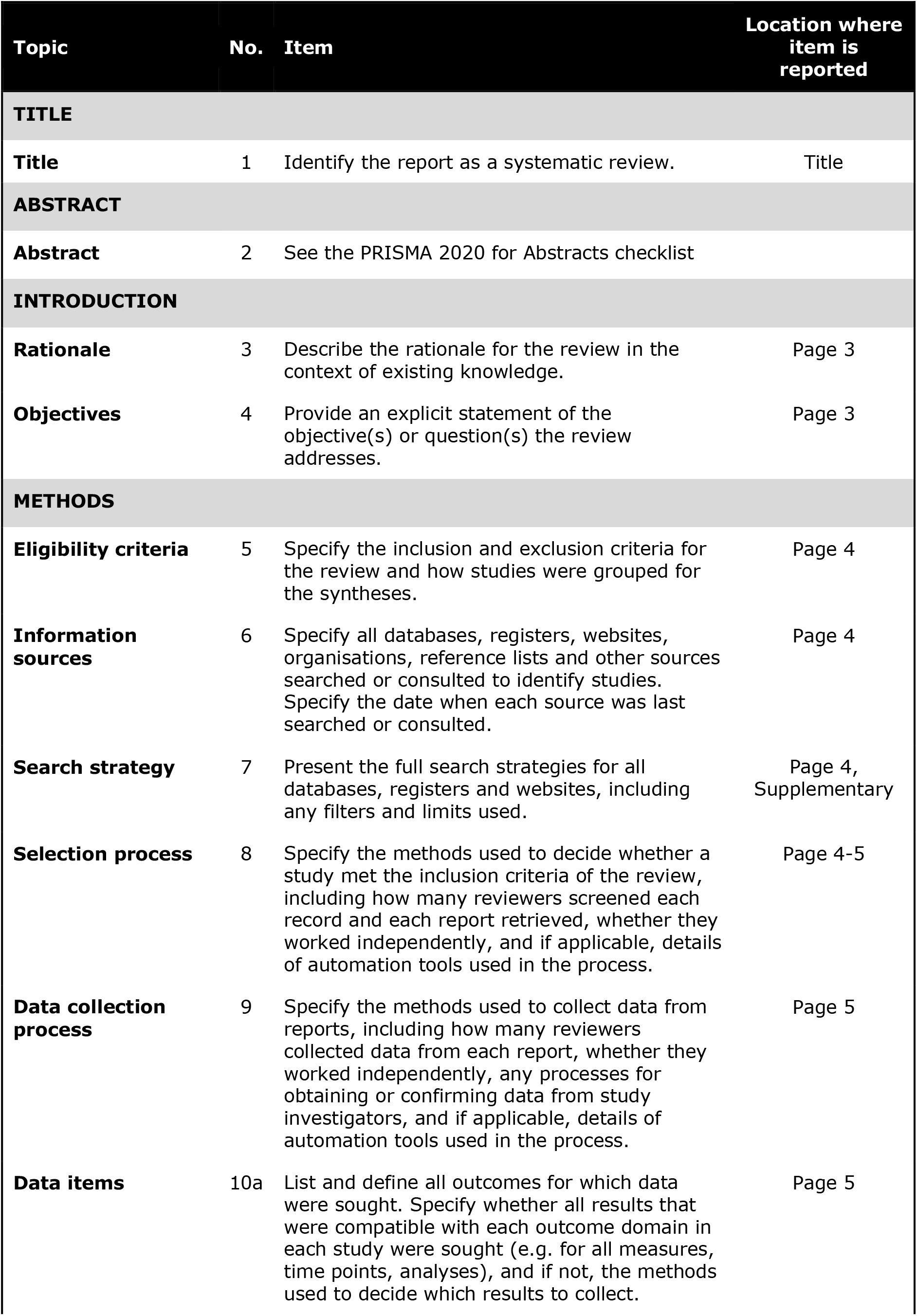
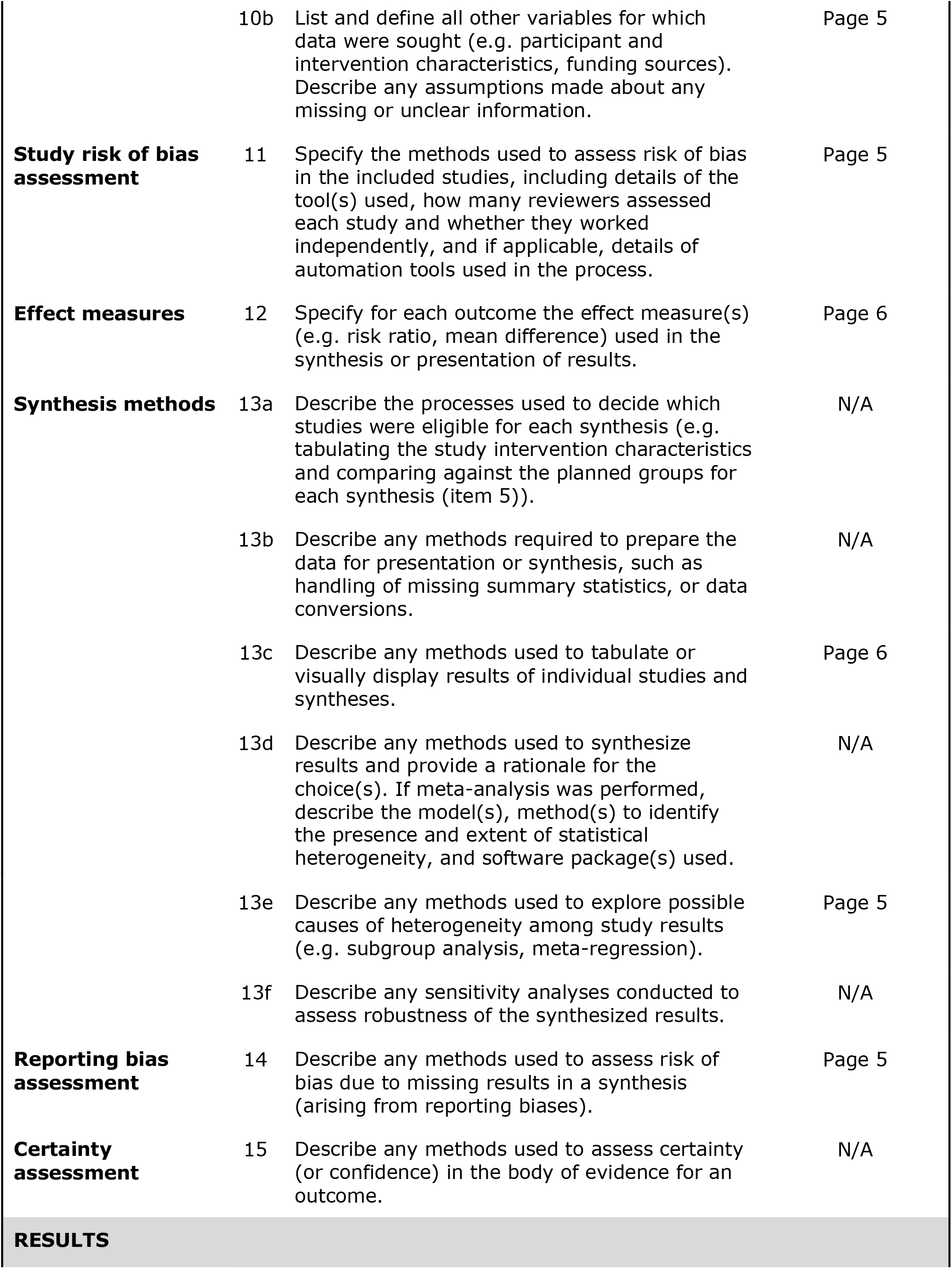
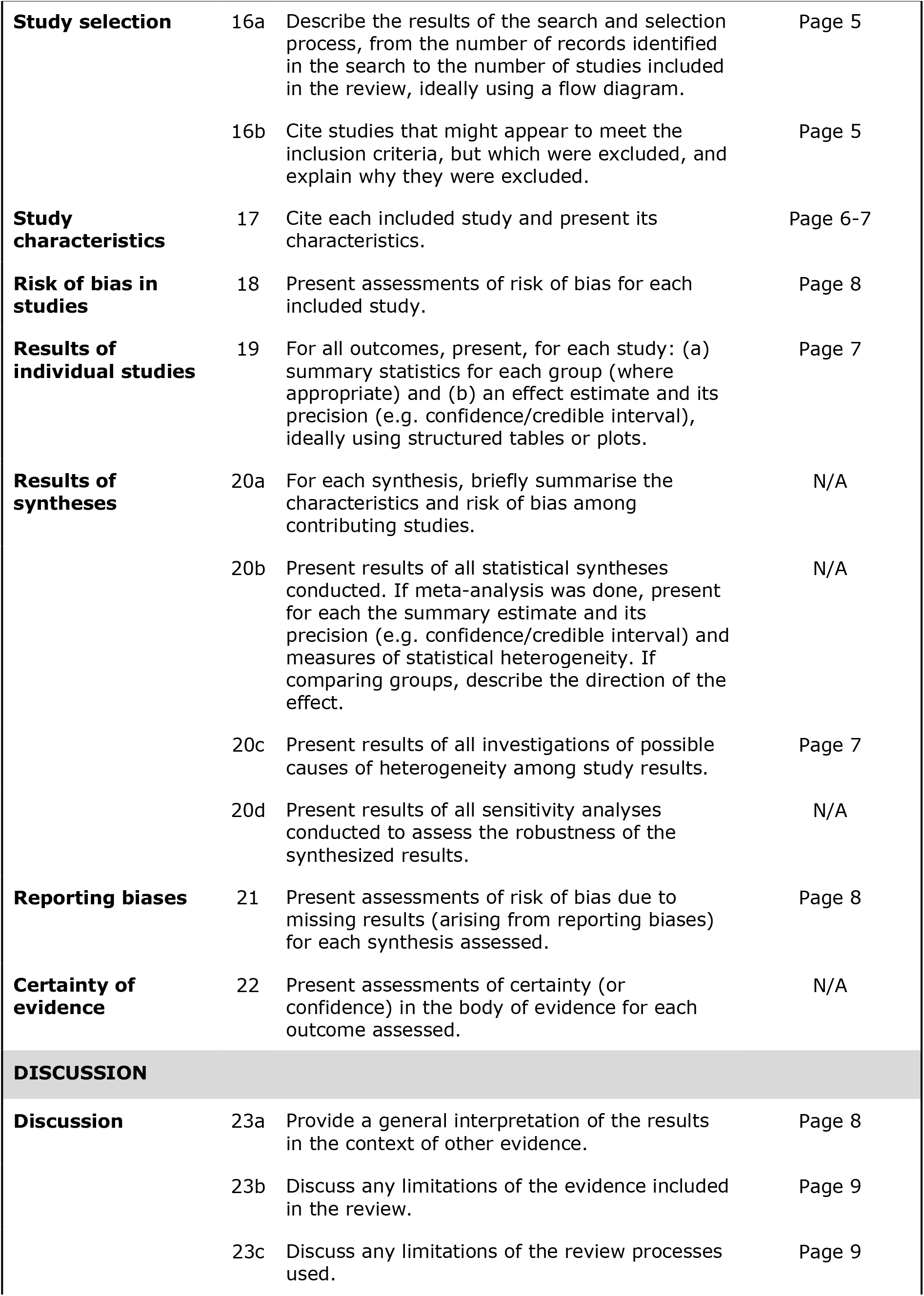
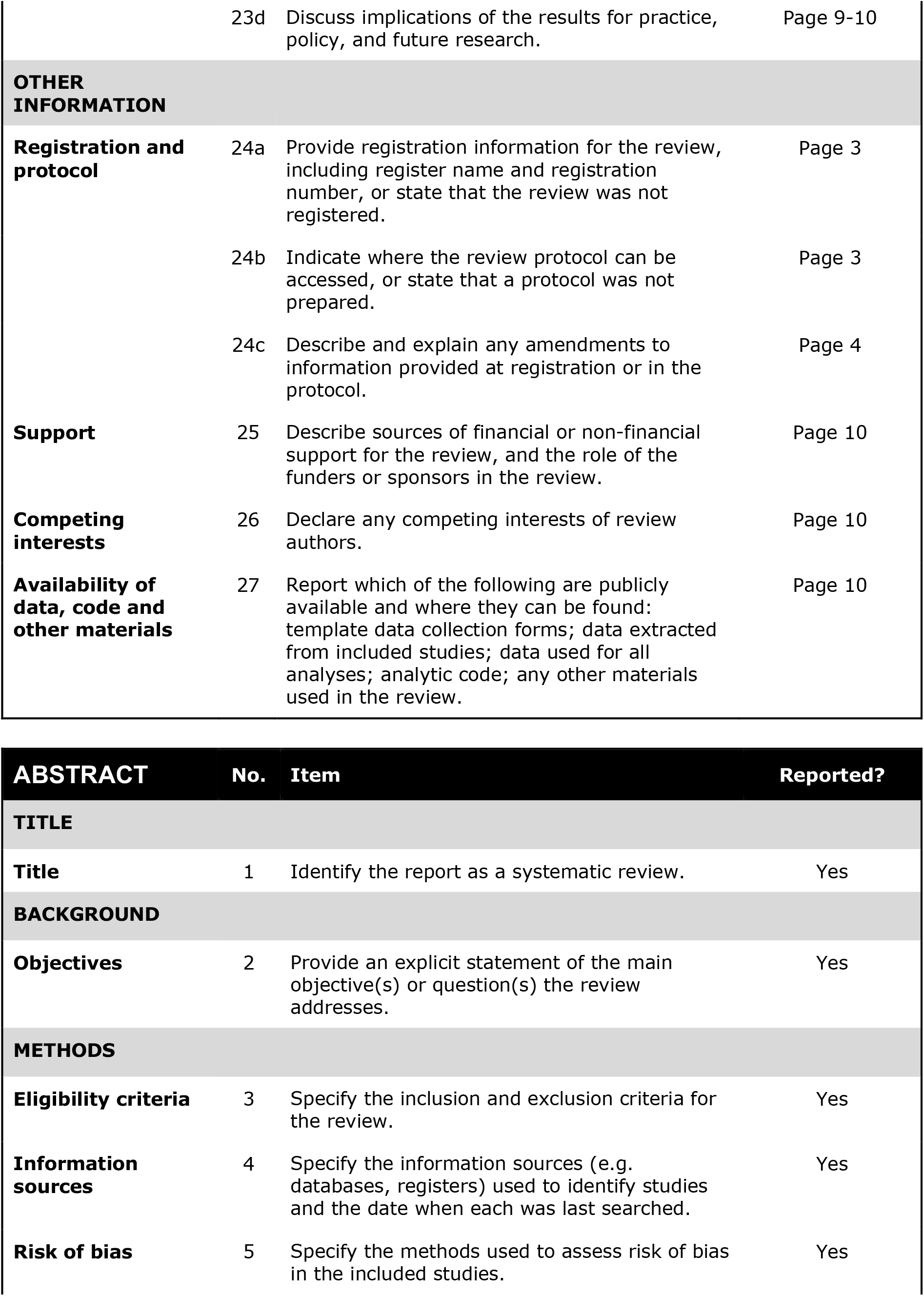
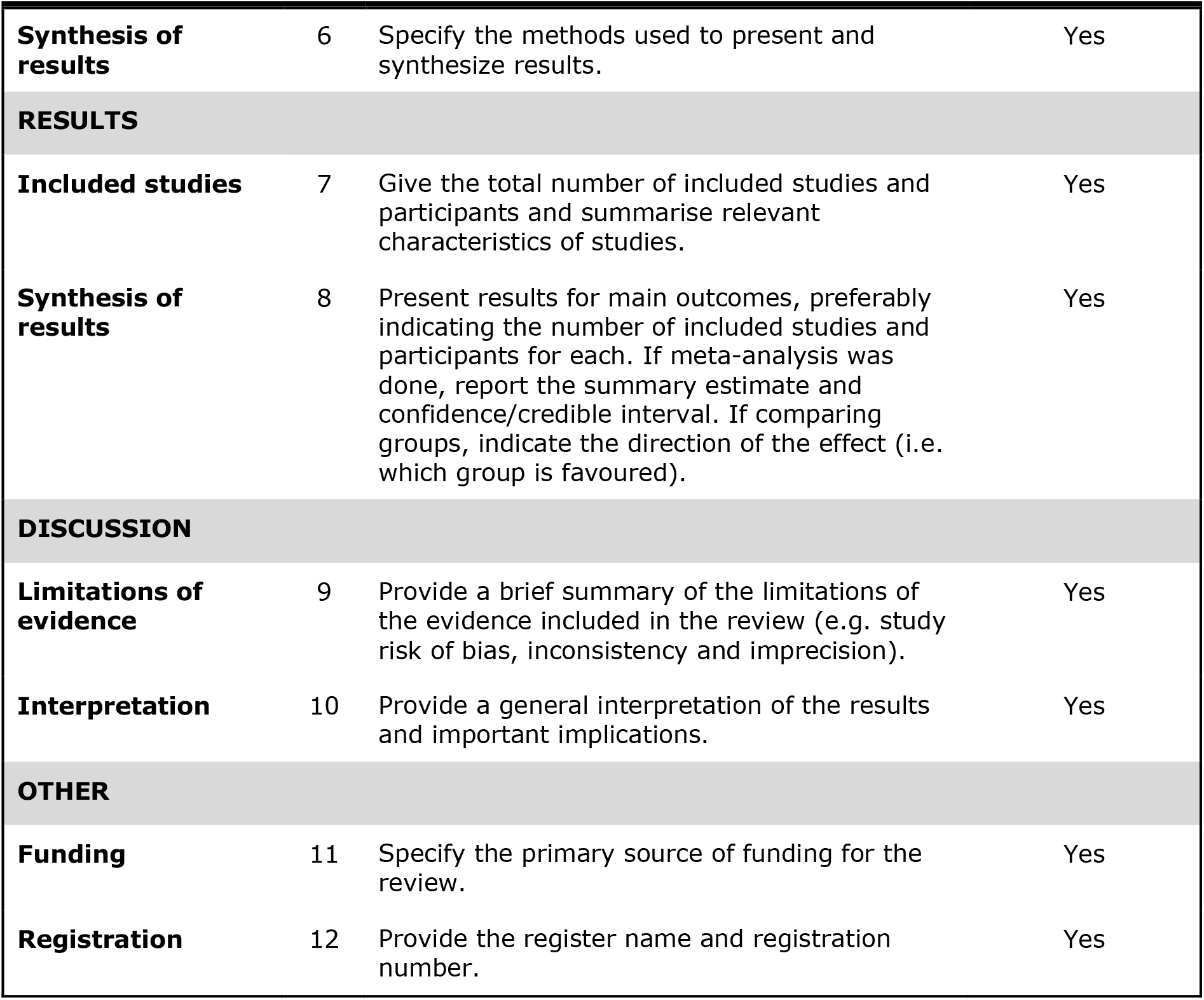
PRISMA checklist for main manuscript and abstract

**S4 Table.** Reasons for exclusion after full text review

| Reason and number excluded | Explanation |
| --- | --- |
| Wrong time period (n=89) | All data or majority of data were gathered prior to 2016. |
| Wrong publication type (n=12) | Article was a protocol, review article or qualitative report without any quantitative descriptors. |
| Wrong population (n=115) | Population for main analysis included participants younger than 18yrs without stratification.<br>Population for main analysis excluded those with ART experience or required a minimum follow-up period after treatment initiation. |
| Not sufficient data for analysis (n=106) | No exclusion criteria met, but data reported was not sufficient for us to determine naïve vs non-naïve status. |

